# Reduction of clonal hematopoiesis mutation burden in coronary patients treated with low-dose colchicine

**DOI:** 10.1101/2024.10.17.24315679

**Authors:** Jean-Claude Tardif, Lambert Busque, Steve Geoffroy, Johanna Sandoval, Louis-Philippe Lemieux Perreault, Ian Mongrain, Diane Valois, Marie-Josée Gaulin-Marion, Manuel Buscarlet, Sylvie Provost, Aldo P. Maggioni, Simon Kouz, Fausto J. Pinto, Jose Lopez-Sendon, David D. Waters, Rafael Diaz, Habib Gamra, Ghassan S. Kiwan, Colin Berry, Wolfgang Koenig, Jean C. Grégoire, Philippe L. L’Allier, Mylène Provencher, Marie-Claude Guertin, François Roubille, Essaid Oussaid, Amina Barhdadi, Marie-Pierre Dubé

## Abstract

Clonal hematopoiesis involves mutations in hematopoietic stem/progenitor cells, which increase the risk of cardiovascular disease, particularly under pro-inflammatory conditions. This study assessed the impact of the anti-inflammatory medication colchicine on clonal hematopoiesis in patients with recent myocardial infarction from the COLCOT trial. Participants were randomly assigned to low-dose colchicine (0.5 mg daily) or placebo, with 848 providing two DNA samples for longitudinal analysis. Targeted error-corrected sequencing was used, and 15,919 mutations were followed over a median period of 19.5 months. The results showed significantly lower variant allele fractions in the colchicine group compared to placebo (p interaction=0.03), with notable reductions in *TET2* (10.3%, p=0.007; p interaction=0.001), *TP53* (11.8%, p=0.001; p interaction=0.03), and *SF3B1* mutations (19.9%, p=0.006; p interaction=0.005). Thus, colchicine reduced the proportion of clonal hematopoiesis mutations in patients with coronary disease, and longer-term studies with diverse populations are needed to confirm its potential benefits in mitigating related health risks.

## INTRODUCTION

Human hematopoietic stem/progenitor cells are responsible for producing more than 10 billion blood cells daily throughout a person’s lifespan. With aging, mutations can occur in these cells conferring a competitive growth advantage, leading to gradual clonal dominance of hematopoiesis.^1,2^ Large cohort studies have demonstrated that clonal hematopoiesis (CH) is associated with an increased risk of developing myeloid malignancies and cardiovascular disease ^3–5^ and may be a biomarker of unhealthy aging. Moreover, CH was shown to be unidirectionally linked to the development of atherosclerosis, suggesting a potential causal role in these pathologies.^6^ The prevalence of CH is a function of age, depth of sequencing, and number of genes analyzed.^7^ The World Health Organisation^8^ distinguishes age-related CH, which has no specific quantitative threshold for mutation burden, from clonal hematopoiesis of indeterminate potential (CHIP)^9^ where the variant allele frequency (VAF) has to be ≥ 2%. Somatic mutations in over 100 genes have been identified as candidate drivers of CH, with the most common mutations being in the three epigenetic regulators: DNA methyltransferase 3a (*DNMT3A*), ten-eleven translocation-2 (*TET2*), and additional sex combs-like 1 (*ASXL1*). Risk factors for adverse events in CHIP carriers include clone size, driver gene identity, and number of mutations.^10^

Carriers of CHIP exhibit a pro-inflammatory response, with higher levels of C-reactive protein, IL-6, IL-1β, IL-18 and TNF-α compared to non-carriers.^11–13^ In a pro-inflammatory environment, mutant hematopoietic stem/progenitor cells display an attenuated inflammatory response relative to wild-type hematopoietic stem/progenitor cells, providing them with a relative fitness advantage over non-mutated cells.^14,15^ The proliferation of these mutated cells can be mitigated by the experimental neutralization of pro-inflammatory interleukin-6 and IL-1 signaling.^16–18^ A post-hoc analysis of the CANTOS trial showed that carriers of CH mutations in the *TET2* gene experienced fewer cardiovascular events when treated with the anti-inflammatory monoclonal antibody canakinumab, which targets interleukin-1.^19^ Inflammation plays an important role in atherosclerosis,^20^ and large randomized clinical trials have shown the benefits of inflammation reduction on cardiovascular outcomes in patients with atherosclerotic cardiovascular disease, including the COLCOT and LoDoCo2 studies with low-dose colchicine.^21–23^

Colchicine is a potent and widely used anti-inflammatory medication that acts by inhibiting tubulin polymerization and microtubule formation, leading to effects on the inflammasome, inflammatory chemokines, and adhesion molecules.^11–13^ In this study, we used error-corrected sequencing to investigate the effect of colchicine compared to placebo on changes in CH variant allele frequency (VAF) over time in patients who were recruited following a recent myocardial infarction as part of the COLCOT study. We then assessed whether individuals with CHIP derived greater benefits in the reduction of clinical events as compared to non-CHIP carriers. The study provides evidence supporting colchicine’s ability to contribute to the reduction of CH mutations in some of the most common and clinically relevant genes involved in CH, *TET2, TP53* and *SF3B1,* possibly intercepting clonal progression and reducing disease risks in carriers. In line with recent findings, our results support the potential for using therapeutic strategies targeting inflammation and inflammasomes to reduce the risk of cardiovascular events in patients with *TET2* CH mutations.^24^

## RESULTS

### PARTICIPANTS

Of the 4745 patients enrolled in the COLCOT trial, 1610 were evaluated for CH. Table 1 presents the baseline characteristics of these patients. The average age of participants was 60.6 years, 18.1% were female, and the mean BMI was 28.8 kg/m^2^. For 25% of the participants, the first DNA sample was collected within 24 days of initiating the study drug. The median number of days between the baseline visit and the first DNA sample collection was 187 (6.1 months). Of the 1610 participants, 29 had only one DNA sample collected at the end-of-study visit. For the 848 participants with two DNA samples, the median interval between the first and second DNA sample was 19.5 months. The primary endpoint in the COLCOT trial consisted of cardiovascular death, resuscitated cardiac arrest, myocardial infarction, stroke, or urgent hospitalization for angina requiring coronary revascularization, to which we added any coronary revascularization event for the purpose of the present study. This composite endpoint occurred in 8.7% and 7.4% of participants in the placebo and colchicine arms respectively in the present study, compared to 9.3% and 7.6% in the placebo and colchicine arms respectively in the main COLCOT trial.

**Table 1.**
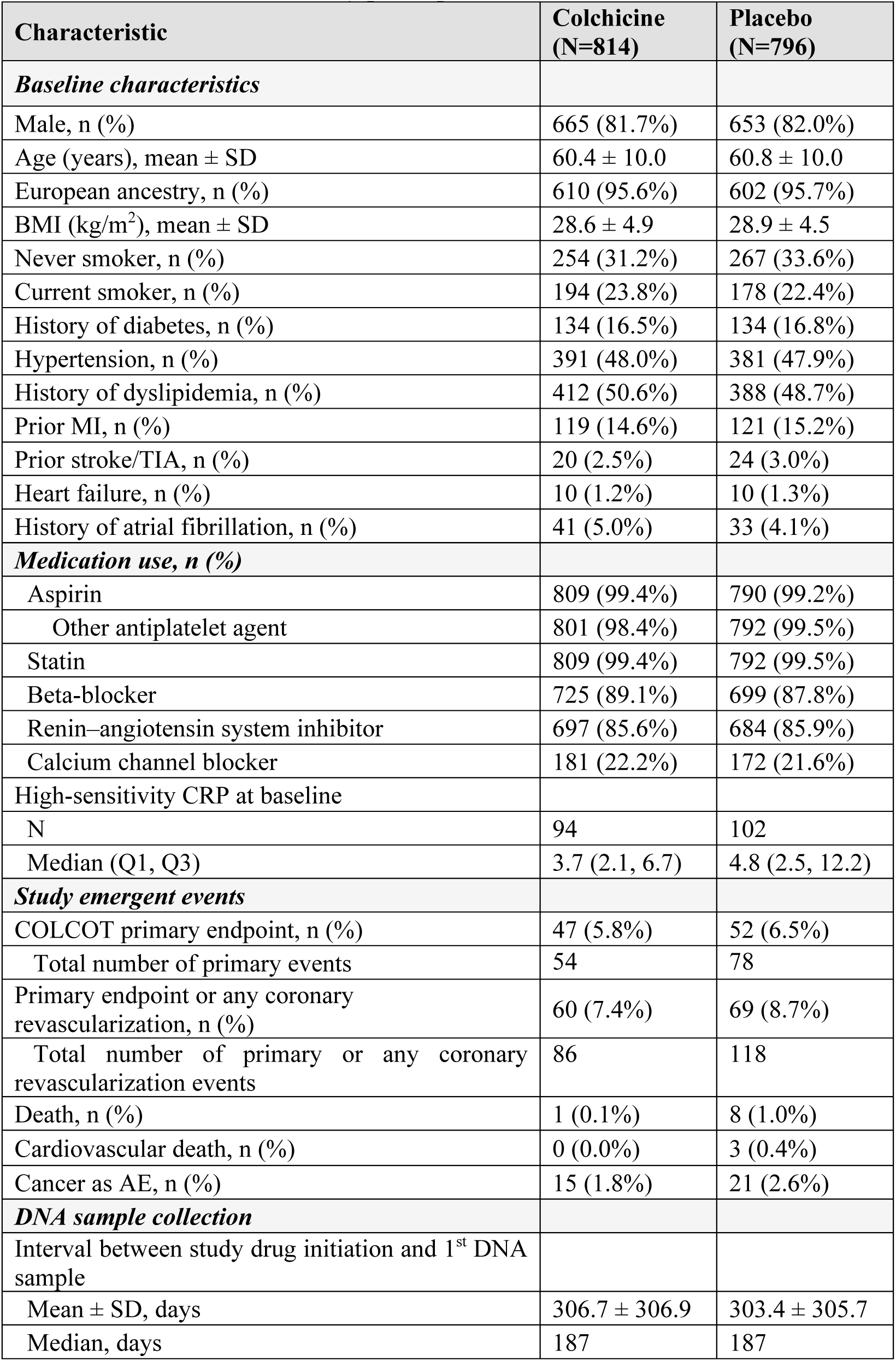
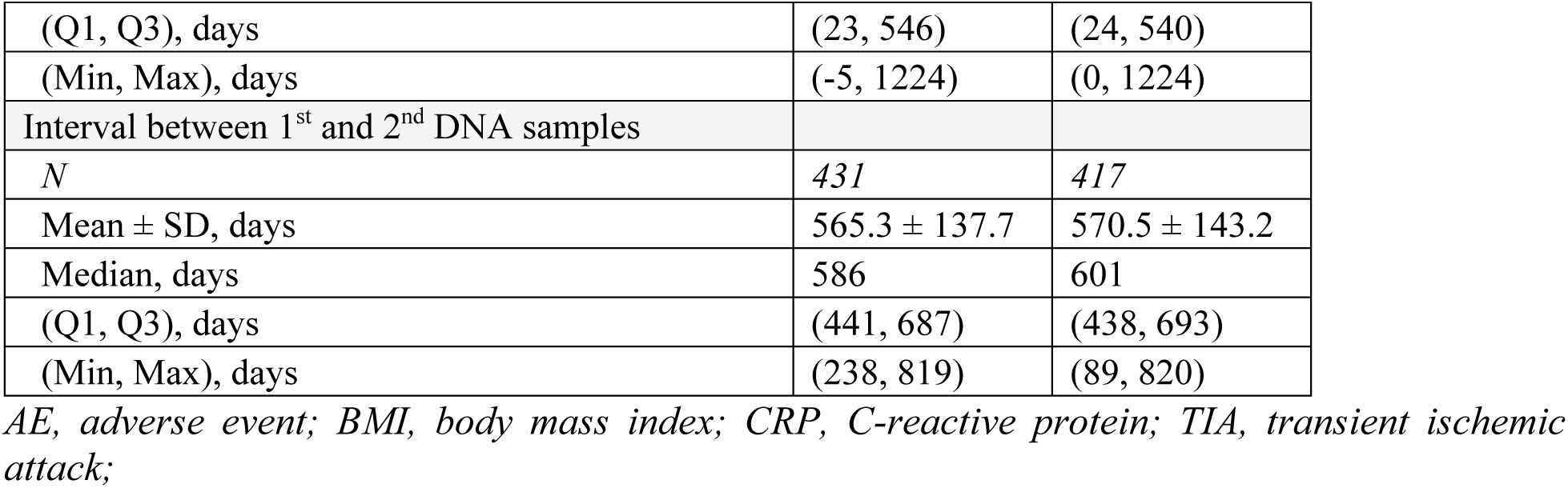
Characteristics of study participants.

### CHANGE IN VAF OF CLONAL HEMATOPOIESIS MUTATIONS

We performed targeted error-corrected deep sequencing of 11 of the most common genes driving clonal expansion. The sequencing libraries were constructed with unique molecular indices to label both DNA strands, enabling the creation of a consensus sequence and enhancing the error detection capability. Sequencing was conducted at an average depth of 64,232x, resulting in an average consensus read depth of 3,896x after accounting for unique molecular tags. When categorizing individuals according to specific VAF groups based on the mutation with the highest VAF per gene, we observed a high prevalence of mutation carriers with low VAF. Specifically, for VAF < 0.5% in the first DNA sample, 77.4% of individuals had *TET2* mutations, 68.7% had *DNMT3A* mutations and 64.8% had *TP53* mutations (Supplementary Table 6a). The prevalence of CH mutations carriers within the 0.5% to 2% VAF range was 14.2% for *TET2*, 12.7% for *DNMT3A* and 2.0% for *TP53*. As expected, the prevalence of mutation carriers for mutations of VAF ≥ 0.5% increased with age (Figure 1b; Supplementary Table 6b). However, for individuals with mutations of VAF < 0.5%, the prevalence of *DNMT3A* and *TET2* mutation carriers significantly decreased with age (p < 0.01), suggesting possible selective clonal dynamics favoring higher VAF mutations in aging individuals.

**Figure 1.**
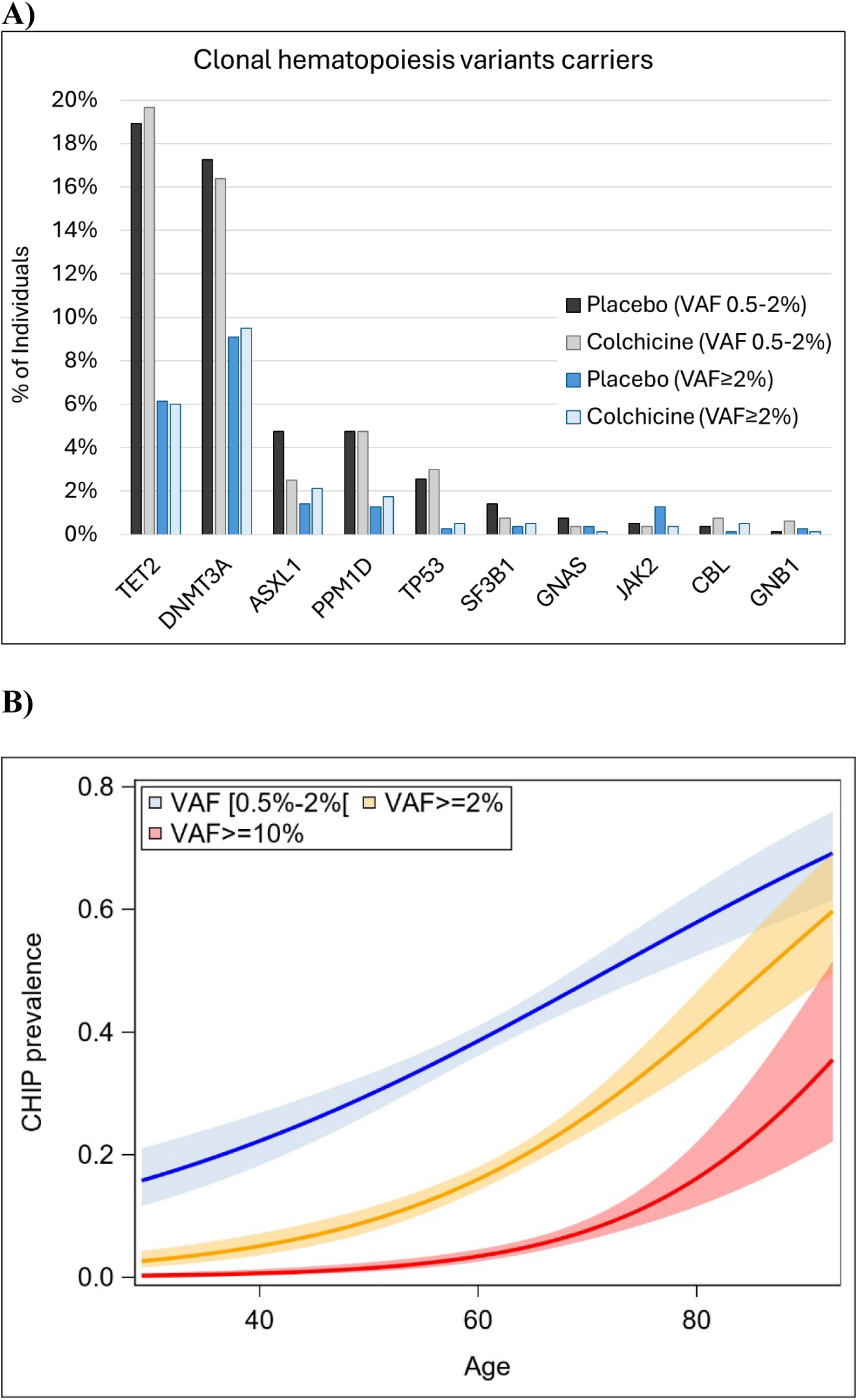
Prevalence of clonal hematopoiesis variant carriers, based on the mutation with the highest variant allele fraction (VAF) value in the first sample collected. **A.** Percentage of carrier individuals by gene, group, and VAF category. **B.** Prevalence of variant carriers of any gene mutation by age.

CHIPs (VAF ≥ 2%) were detected in 294 out of 1610 participants (18.3%) across the 11 tested genes, considering their occurrence at either the first or second DNA samples (Figure 1a; Supplementary Table 6b). The most frequently observed CHIP genes were *DNMT3A* (9.1%), *TET2* (6.0%) and *ASXL1* (1.7%), consistent with previous studies.^13^

In the subgroup of 848 individuals with two timepoint samples, we identified 1954 unique CH variants among 15,919 longitudinal observations (Supplementary Table 7a). There were 7,878 longitudinally assessed mutations in the placebo group, with an observed mean increase in VAF of 0.00011, and 8,041 mutations in the colchicine group with an observed mean increase in VAF of 0.00007. We compared the VAFs of mutations at the end-of-study sample to that of the first sample by using a generalized mixed model that fits a beta-binomial distribution with a logit link function. We found that the VAFs of mutations at the end-of-study samples in the colchicine group were significantly lower compared to those of the placebo group (p_int_ = 0.03). Notably, the VAF of *TET2* variants increased by 9.1% in the end-of-study samples compared to the first sample in the placebo group (p = 0.05) whereas the VAF of *TET2* variants decreased by 10.3% in the colchicine group (p = 0.007), with a significant difference between colchicine and placebo groups (p_int_ = 0.001) (Table 2). We also observed significant reductions of 11.8% in the VAF of *TP53* mutations (p = 0.001; p_int_ = 0.03) and 19.9% in the VAF of *SF3B1* mutations (p = 0.006; p_int_ = 0.005) with colchicine (Table 2 and Supplementary Table 7a). Surprisingly, a significant reduction in VAFs for *JAK2* mutations was observed in the placebo group (−28.1%, p = 3.4×10^-5^) but not in the colchicine group (P_int_ = 0.003). However, this difference was largely due to patients with VAF≥ 2% (n=5/185) with only 1 patient in the colchicine group. This difference was not seen in smaller clones.

**Table 2.** Results of the analysis of change in clonal hematopoiesis variant allele frequency between two sample timepoints.

| Gene | Group | N unique patients | Pairwise observations | Mean observed VAF difference | Estimate (SE)* | % change in VAF | p value | Interaction p value |
| --- | --- | --- | --- | --- | --- | --- | --- | --- |
| Any gene | Colchicine | 431 | 8,041 | 0.00007 | -0.055 (0.043) | -5.4% | 0.203 | 0.033 |
|  | Placebo | 417 | 7,878 | 0.00011 | 0.079 (0.045) | 8.2% | 0.081 |  |
| *ASXL1* | Colchicine | 335 | 715 | 0.00002 | 0.110 (0.057) | 11.6% | 0.052 | 0.464 |
|  | Placebo | 338 | 673 | -0.00003 | 0.029 (0.081) | 3.0% | 0.718 |  |
| *CBL* | Colchicine | 32 | 37 | 0.00186 | 0.217 (0.131) | 24.3% | 0.132 | 0.456 |
|  | Placebo | 29 | 29 | -0.00007 | 0.079 (0.117) | 8.2% | 0.535 |  |
| *DNMT3A* | Colchicine | 420 | 1,802 | 0.00012 | 0.043 (0.065) | 4.4% | 0.505 | 0.132 |
|  | Placebo | 405 | 1,785 | 0.00017 | 0.185 (0.066) | 20.3% | 0.005 |  |
| *GNAS* | Colchicine | 81 | 86 | 0.00001 | -0.171 (0.078) | -15.7% | 0.048 | 0.303 |
| | Placebo | 85 | 93 | 0.00012 | -0.285 (0.051) | -24.8% | $3.39 \times 10^{-5}$ | |
| *GNB1* | Colchicine | 15 | 15 | 0.00083 | 0.173 (0.093) | 18.8% | 0.124 | 0.065 |
|  | Placebo | 12 | 12 | -0.00147 | -0.180 (0.075) | -16.4% | 0.096 |  |
| *JAK2* | Colchicine | 77 | 86 | -0.00022 | 0.167 (0.137) | 18.2% | 0.235 | 0.003 |
| | Placebo | 85 | 99 | -0.00065 | -0.330 (0.086) | -28.1% | $7.68 \times 10^{-4}$ | |
| *PPM1D* | Colchicine | 102 | 122 | 0.00110 | 0.274 (0.061) | 31.4% | $2.75 \times 10^{-5}$ | 0.140 |
|  | Placebo | 107 | 128 | 0.00015 | 0.125 (0.070) | 13.3% | 0.077 |  |
| *SF3B1* | Colchicine | 174 | 216 | 0.00003 | -0.223 (0.078) | -19.9% | 0.006 | 0.005 |
|  | Placebo | 163 | 192 | 0.00011 | 0.130 (0.098) | 13.9% | 0.188 |  |
| *TET2* | Colchicine | 431 | 4,108 | 0.00004 | -0.109 (0.041) | -10.3% | 0.007 | 0.001 |
|  | Placebo | 416 | 4,029 | 0.00014 | 0.087 (0.045) | 9.1% | 0.055 |  |
| *TP53* | Colchicine | 364 | 854 | -0.00002 | -0.125 (0.038) | -11.8% | $9.82 \times 10^{-4}$ | 0.026 |
|  | Placebo | 353 | 837 | 0.00004 | 0.019 (0.048) | 1.9% | 0.699 |  |

\* Generalized mixed model fitting a beta binomial with logit link function for the VAF defined as VD/DP, adjusted for the interval in days between sample collections and patient age. The reported estimate is for the timepoint variable contrasting end-of-study to 1st sample. The % change in VAF was obtained from the ratio of the estimated VAF at end-of-study over the estimated VAF in the 1<sup>st</sup> sample.
DP, sequencing read depth; SE, standard error; VAF, variant allele fraction; VD, variant counts.

The analysis of patients with CHIP (VAF ≥ 2%) was limited by the smaller number of observations, which was 100 times less than the overall cohort, and did not reach statistical significance when comparing the placebo and colchicine groups (Supplementary Table 7a). However, we observed that clones grew more rapidly in the placebo group when restricting to VAFs ≥ 2% compared to no restriction (15.5% vs. 8.2%), suggesting that clonal expansion may be more pronounced as clones reach a certain proportion of hematopoiesis.

In sensitivity analyses, we considered all variants observed without restriction on VD (VD ≥ 1). While lowering the VD threshold from 5 to 1 increases the number of false positive variants, it also increases the total number of observations detected, including true positive mutations, and improves the detection of mutations with very low VAF (Supplementary Table 5). With a VD threshold of 1, we included 206,015 pairwise observations into the analysis. Using a mixed regression model fitting a binomial negative distribution to account for the predominance of null VAF, the results obtained were consistent with those obtained at the threshold of VD ≥ 5. Over all CH mutations, we found a significant difference in the counts of mutant variants between the colchicine and placebo groups (p_int_=8.0×10^-4^), with an increase in counts of 5.3% in the placebo group (p=1.1×10^-11^) compared to an increase in counts of 1.8% in colchicine group (p=0.009) (Supplementary Table 7b). Notably, for *TET2* mutations, an increase in counts of 5.7% was observed in the placebo group (p=1.2×10^-10^) compared to an increase of 2.1% in the colchicine group (p=0.01; p_int_=0.003).

### IMPACT ON CARDIOVASCULAR ENDPOINTS

In patients with *TET2* CHIP mutations (VAF ≥ 2%), the composite endpoint occurred in 2 (4.2%) patients in the colchicine group and 4 (8.3%) patients in the placebo group (hazard ratio (HR) = 0.62; 95% confidence interval (CI): 0.11-3.42; p = 0.58). Among patients without *TET2* CHIP mutations, the composite endpoint occurred in 58 (7.6%) patients in the colchicine group and 65 (8.7%) patients in the placebo group (HR = 0.85; 95% CI: 0.60-1.21; p = 0.36). Figure 2 illustrates the cumulative incidence curves for the composite endpoint. For patients with *TET2* CHIP mutations, the total number of endpoint events (first and subsequent) was 2 in the colchicine group and 5 in the placebo (HR = 0.49; 95% CI: 0.09-2.83; p = 0.43, Table 3). In patients without *TET2* CHIP mutations, the total number of events was 84 in the colchicine and 113 in the placebo group (HR = 0.71; 95% CI: 0.48-1.05; p = 0.09, Table 3). Sensitivity analyses of *TET2* mutation carriers at VAF ≥ 0.5% and ≥ 1% were concordant, showing lower event rates in *TET2* mutation carriers compared to non-*TET2* carriers and to individuals without CH mutations, although these differences did not reach statistical significance (Supplementary Table 8a, b). The low number of carriers of *TP53* and *SF3B1* mutations precluded the conduct of stratified analyses with the composite endpoint at VAF ≥ 0.5%. For VAF > 0, when testing for the reduction in composite endpoint events with colchicine compared to placebo in mutation carriers and non-carriers of *TP53* and *SF3B1* mutation carriers, the difference did not reach statistical significance (Supplementary Table 8a, b).

**Figure 2.**
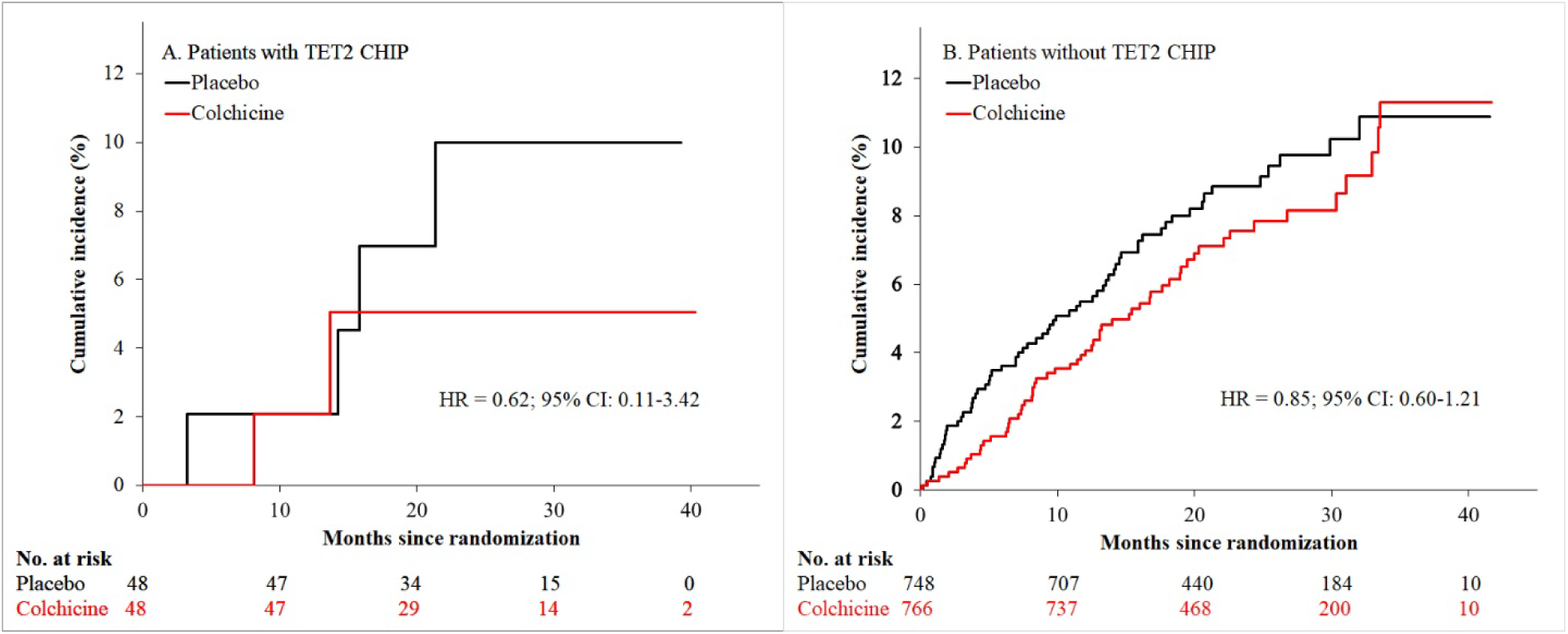
Cumulative incidence curves of the cardiovascular composite endpoint (cardiovascular death, resuscitated cardiac arrest, myocardial infarction, stroke, urgent hospitalization for angina requiring coronary revascularization, or any coronary revascularization) in the colchicine group (red) and the placebo group (black), in a time-to-event analysis, for patients **A.** with *TET2* CHIP mutations and **B.** without *TET2* CHIP mutations.

**Table 3.** Hazards ratios of the effect of colchicine versus placebo on cardiovascular clinical events in patients with *TET2* CHIP mutations.

| Clinical Outcomes<sup>†</sup> | Patients | N | Colchicine | Placebo | HR (95% CI)<sup>‡</sup> | P |
| --- | --- | --- | --- | --- | --- | --- |
|  |  |  | N patients with events |  |  | Value |
| Composite endpoint (time to first event) | With *TET2* CHIP | 96 | 2 (4.2%) | 4 (8.3%) | 0.62 (0.11-3.42) | 0.58 |
|  | Without *TET2* CHIP | 1514 | 58 (7.6%) | 65 (8.7%) | 0.85 (0.60-1.21) | 0.36 |
|  | Without any CHIP | 1333 | 53 (8.0%) | 60 (9.1%) | 0.85 (0.59-1.23) | 0.39 |
| Recurrent event analysis |  |  | N total events |  | HR (95% CI)<sup>¶</sup> |  |
| Total number of endpoint events (first and subsequent) | With *TET2* CHIP | 96 | 2 | 5 | 0.49 (0.09, 2.83) | 0.43 |
|  | Without *TET2* CHIP | 1514 | 84 | 113 | 0.71 (0.48, 1.05) | 0.09 |
|  | Without any CHIP | 1333 | 75 | 104 | 0.70 (0.47, 1.06) | 0.09 |

CHIP, clonal hematopoiesis of indeterminate potential; HR, hazard ratio. <sup>†</sup>The composite primary endpoint is composed of cardiovascular death, resuscitated cardiac arrest, myocardial infarction, stroke, urgent hospitalization for angina requiring coronary revascularization, or any coronary revascularization. <sup>‡</sup>Cox proportional hazards model adjusted for age and sex. <sup>¶</sup> Andersen and Gill (AG) model adjusted for age and sex.

## DISCUSSION

CH has emerged as a candidate biomarker of disease prediction and progression, with a complex relationship with the body’s inflammatory response mechanisms. In this study, we explored the impact of colchicine, a potent anti-inflammatory medication, on CH. By using error-corrected targeted sequencing with unique molecular tags at a mean sequencing depth of 64,232x, we reached a high level of precision that enabled the detection of subtle changes in variant allele fraction (VAF) that are typically missed with whole exome or whole genome sequencing. We observed significant changes in the proportion of mutant cells over a median period of 1.5 years in individuals randomly assigned to receive low-dose colchicine (0.5 mg once daily) or placebo. Our results show that over this period, individuals in the colchicine group had significantly lower clonal expansion as compared to those in the placebo group. When broken down into specific genes, we found that the proportion of mutant cells was significantly decreased for *TET2, TP53*, and *SF3B1* mutations in the colchicine group, whereas the proportion increased in the placebo group, with a significant difference between the two groups. Specifically, in the colchicine group, the VAF of *TET2* mutations decreased by 10%, that of *TP53* mutations decreased by 12% and the VAF of *SF3B1* mutations decreased by 20%. These results suggest that colchicine can attenuate the expansion of these clones, possibly by modulating the inflammatory environment.

Colchicine’s inhibition of tubulin polymerization and subsequent disruption of microtubule formation leads to reduced neutrophil activity and suppression of the NLRP3 inflammasome.^25^ This mechanism effectively decreases the production of pro-inflammatory cytokines such as IL-1β and IL-18, which are known to foster a pro-inflammatory environment favoring the expansion of clonal hematopoietic cells. Whether the anti-mitotic effects of colchicine also contribute to the reduced progression of hematopoietic clones cannot be excluded, however its more marked impact on clones with lower VAFs makes this mechanism less probable. Our results suggest that by reducing the inflammatory milieu, colchicine may reduce the selective advantage of mutated hematopoietic stem/progenitor cells. Previous studies have highlighted the role of inflammation on the progression of CH and its associated risks, particularly for *TET2* mutations, which were shown to thrive in pro-inflammatory conditions.^14,15^. Individuals with CH exhibit elevated levels of inflammatory markers, and those with *TET2* mutations in particular demonstrate an increased production of IL-6, IL-1β, IL-18 and TNF-α.^4,11–13^ Inhibiting IL-1β with the monoclonal antibody canakinumab in the CANTOS trial was shown to reduce cardiovascular events in patients with *TET2* CH, emphasizing the relevance of inflammation control for CH-related clinical outcomes.^19^ In our analysis, we assessed the risk of cardiovascular events in patients with and without *TET2* mutations who were randomly assigned to receive low-dose colchicine or placebo. The cardioprotective effect of colchicine as compared to placebo appeared to be greater among the patients with *TET2* mutations compared to those without any CH mutations, however, these differences did not reach statistical significance. The inability to reproduce the clinical impact of therapeutic modulation of inflammation in CHIP carriers seen in the CANTOS trial may be due to the lower rate of cardiovascular events in COLCOT, to differences in patient populations, treatment protocols, the difference in follow-up duration, or to differences in variant filtering algorithms. Nonetheless, our study suggests that colchicine’s broad anti-inflammatory effects, which target multiple inflammatory pathways, might offer a valuable approach for the management of CH mutation expansion and its related outcomes for *TET2, TP53* and *SF3B1* mutations.

TP53 is crucial in maintaining genomic stability and regulating cell proliferation and apoptosis. In hematopoietic stem/progenitor cells, *TP53* mutations can disrupt these regulatory mechanisms, leading to uncontrolled cell growth and an increased likelihood of clonal expansion. This is particularly relevant in an inflammatory environment, which can induce DNA damage and increase the selective pressure for cells with *TP53* mutations, as these cells are more likely to survive and proliferate despite the cellular stress.^26^ In conditions such as myelodysplastic syndromes and acute myeloid leukemia, *TP53* mutations are often linked to poorer prognosis.^27^ The inflammatory milieu further exacerbates the selective advantage of *TP53*-mutated hematopoietic stem/progenitor cells by creating a setting where these cells can outcompete their normal counterparts, contributing to disease progression and resistance to therapy.^28^ *TP53*-mutated cells are actively selected under genotoxic exposure such as chemotherapy or radiotherapy and are linked to secondary myeloid cancers.^29^ In our study, the high analytic sensitivity allowed the observation that 85% of the cohort population had a *TP53* mutation present mainly at low VAF (<0.5%). *TP53* mutations were significantly less likely to expand in the colchicine group compared to the placebo group. It is possible that colchicine’s inhibition of the NLRP3 inflammasome and associated pro-inflammatory cytokines may diminish the selective advantage of *TP53* mutated cells. Our findings suggest that colchicine may be beneficial in managing clonal expansions driven by *TP53* mutations, potentially lowering the risk of progression to more aggressive hematological conditions and preventing secondary cancer in selected individuals at risk. Similarly, the likelihood of clonal expansion of *SF3B1* mutations were significantly lowered with colchicine in our study, highlighting another important aspect of CH. *SF3B1* encodes a core component of the spliceosome which is crucial for the splicing of pre-mRNA, and consequentially, *SF3B1* mutations lead to the production of aberrant splice variants. *SF3B1* mutations can dysregulate various cellular functions and pathways, including heme biosynthesis, mitochondrial metabolism, and the NF-κB pathway.^30,31^ *SF3B1* mutations in myelodysplastic syndromes patients were shown to enhance proinflammatory gene expression in blast cells, possibly fostering a pro-inflammatory milieu.^32^ The detection of *SF3B1* mutations in the context of CH is clinically relevant as it may influence risk assessment and therapeutic strategies to prevent progression to malignancies.^33^ Our findings suggest that colchicine may help reduce clonal expansion of *SF3B1* mutations.

Our study has certain limitations. First, only 1610 patients participated in the CH substudy of COLCOT, with 848 of them providing two DNA samples for assessments of changes in mutant cell fractions over the study period. This may introduce selection bias, as participation in this substudy was voluntary and the provision of a second sample related to a healthy survival, potentially biasing our results toward the null. Furthermore, the majority of participants provided their first blood sample several weeks after initiating study treatment. This limits our ability to measure the full impact of colchicine on clonal expansion from the outset of treatment. Future studies should aim to collect blood samples at baseline, prior to drug initiation, to better assess the complete effect of the intervention. In addition, the median follow-up period of 1.5 years may not be sufficient to observe the long-term effects of colchicine on CH and cardiovascular outcomes. CH and its associated risks may evolve slowly, requiring extended follow-up time to detect significant changes. For example, in established myeloproliferative neoplasm, several years of treatment are needed to observe clonal reduction of *JAK2* V617F mutations with either interferon or Jak2 inhibitors.^34,35^ Future studies with longer follow-up periods like COLCOT-T2D are needed to provide a more comprehensive understanding of colchicine’s long-term impact on CH and related diseases. Importantly, the study’s findings specifically pertain to colchicine and may not be generalizable to other anti-inflammatory medications due to differences in their mechanisms of action and potential effects on CH.

Despite these limitations, our study had several notable strengths. The longitudinal design, conducted within the context of a randomized clinical trial, allowed for robust assessment of changes in CH over time under different exposures, a feature that had not yet been evaluated in this context before. By focussing on 11 of the most commonly mutated genes linked to CH, the study provides a thorough assessment of mutations that are likely to be clinically meaningful for patient populations. Furthermore, the use of high-resolution, error-corrected targeted sequencing with unique molecular tags ensured high accuracy and precision in detecting somatic mutations, enhancing the reliability of our findings, especially in a longitudinal context where the detection of small changes is critical. Finally, the statistical approach employed, using a beta-binomial distribution to model the proportion of variant reads, effectively accounted for the inherent variability in read depth across different genomic positions, thereby further enhancing the precision and reliability of our estimates of VAF and their changes over time.

In conclusion, our study demonstrates that in patients with a recent coronary event treated with a low dose of colchicine daily, the proportion of CH mutations in the *TET2*, *TP53* and *SF3B1* genes significantly decreased over the study period, and this effect was not observed in patients in the placebo group. As CH is a risk factor for several diseases, these results have potentially far-reaching implications beyond cardiovascular populations. Our results support the establishment of clinical trials with colchicine in carriers of CH mutations at risk of adverse outcomes, including cardiovascular endpoints and those related to the new onset and progression of hematological cancers.

## ONLINE METHODS

### STUDY DESIGN AND POPULATION

COLCOT was a randomized, double-blind, placebo-controlled, multinational clinical trial of 4745 patients who were randomly assigned to receive either colchicine 0.5 mg once daily or placebo in a 1:1 ratio on top of standard of care and followed for a median of 22.6 months (NCT02551094). The COLCOT trial design and main clinical study results have previously been published.^22^ Briefly, adult patients were eligible if they had experienced a myocardial infarction within 30 days before enrollment, had completed any planned percutaneous revascularization procedures, and were treated according to guidelines that included the intensive use of statins. Patients were excluded if they had severe heart failure, a left ventricular ejection fraction less than 35%, stroke within the past 3 months, coronary bypass surgery within the past 3 years or planned, a history of non-cutaneous cancer within the last 3 years, significant non-transient hematological abnormalities, severe renal disease with serum creatinine greater than twice the upper limit of normal, or severe hepatic disease. Clinical evaluations occurred at 1 and 3 months following randomization and every 3 months thereafter. Potential study endpoints were adjudicated by an independent clinical endpoint committee who were unaware of the trial-group assignments. The COLCOT primary endpoint was a composite of cardiovascular death, resuscitated cardiac arrest, myocardial infarction, stroke, or urgent hospitalization for angina requiring coronary revascularization in a time-to-event analysis. In the present study, we added any coronary revascularization to the composite clinical endpoint to increase event occurrence and improve statistical power.

The genetic study was optional and 53 sites from 7 countries participated, compared to 167 sites from 12 countries in the main COLCOT trial. Blood samples were centrally collected and processed at the Pharmacogenomics Centre in Montreal as previously described.^36^ An initial blood draw was performed at the time of enrollment or a subsequent visit after randomization, and a second blood draw was performed at the end-of-study visit in a subset of patients. 1656 participants consented to take part in the substudy, exclusions included poor DNA quality and technological failures, leaving 1610 patients for analysis (Supplementary Figure 1). Informed consent was obtained from all study participants. The study was approved by the Montreal Heart Institute research ethics committee and complies with the Declaration of Helsinki.

### LIBRARY PREPARATION AND TARGETED SEQUENCING

All samples were sequenced using a custom designed targeted assay using multiplex PCR-based enrichment by CleanPlex technology (Paragon Genomics, Inc., Hayward, CA, USA). The panel captures the exonic regions of 11 of the most commonly mutated genes linked to CH (*ASXL1*, *CBL*, *DNMT3A*, *GNAS*, *GNB1*, *JAK2*, *PPM1D*, *SF3B1*, *SRSF2*, *TET2* and *TP53*), covering 35,840 bases (97.64%) of the coding regions using 322 amplicons (Supplementary Tables 1a-b). The assay covers the full exonic regions of 6 genes (*PPM1D, TP53, SF3B1, TET2, GNB1* and *JAK2*) and partial exonic regions of 5 genes (*ASXL1_*cdr_1, *CBL_*cdr_1*, DNMT3A_*cdr_6*, DNMT3A_*cdr_7 *GNAS_*cdr_3, and *SRSF2_*cdr_1). Genomic DNA was isolated from 5-10 mL of frozen whole blood (Autopure LS V5.0) and quantified by fluorescence (Tecan Infinite F200 Plate Reader). Libraries were prepared from 60 ng of genomic DNA following recommendations of the CleanPlex protocol. A multiplex PCR reaction was performed using unique molecular indices (UMI)-labeled target-specific primers to barcode and amplify the targeted regions. This was followed by post-barcoding purification (CleanMag Magnetic Beads) and an enzymatic background cleaning step was used to remove non-specific PCR product (primer dimers and PCR artifacts), followed by a bead purification. Unique dual indexes (UDI) were used in a final PCR amplification to generate unique libraries per sample. Samples were bead-purified and the integrity, quality and quantity was verified using the TapeStation 4200 D1000 assay. Each library was normalized to 4 nM, pooled and sequenced on Illumina MiSeq Sequencer as a quality control check prior to sequencing. Five sequencing runs were performed on the Illumina NovaSeq 6000 instrument (Illumina Inc., San Diego, CA, USA) using the NovaSeq Control Software (Recipe Fragment v1.7.5, Analysis viewer v2.4.7) with paired-end sequencing 2×150 bp (NovaSeq S4 Reagent Kit v1.5 and NovaSeq XP 4-Lane Kit v1.5) according to the manufacturer’s instructions.

We implemented the recommended Paragon UMI 12 step bioinformatics analysis pipeline. After demultiplexing the sequencing data, the trailing sequencing adapters in the R1 and R2 reads were trimmed using *cutadapt* v.4.6. Trimmed fastq reads were then converted to unmapped alignment (BAM) files. The 16 bp UMI were removed from BAM files to be stored as an alignment tag. Template reads were reconverted to FASTQ format and mapped against the human genome reference GRCh37 using *bwa-mem* v.0.7.17. The mapped and unmapped tagged reads were merged and the alignment file was created. The resulting alignments were processed using the Fulcrum Genomics consensus reads calling workflow (*fgbio* v2.2.1).^37^ Aligned reads were grouped as read families presumed to originate from the same molecule by adjacency and allowing a maximumn of 1 edit between UMIs and keeping only bases with mapping quality ≥ 20. The CallMolecularConsensusReads from *fgbio* was used to create consensus reads based on ≥ 3 identical UMI reads to produce a consensus read. The consensus reads were aligned against the human genome reference GRCh37 using *bwa-mem* v.0.7.17 again to improve alignment accuracy. We implemented the bioinformatics workflow in GenPipes^38^ as per Paragon’s instructions.

### VARIANT CALLING, ANNOTATION AND FILTERING

Variant calling was performed using *vardict* (v4.2.6.1)^39^ using the default cuttoffs to all but the following arguments: -a, meaning amplicon mode, where reads that do not map to the amplicon are skipped. Read pairs were assigned to an amplicon with a minimal overlap fraction of 95% and the PHRED score threshold was set to 20. Variant consequence prediction was performed using Annovar (v.2020-06-08)^40^ and variants were annotated using the refGene hg19 assembly reference and COSMIC v70 hg19 resource^41^. Variants were identified as putative drivers of CH according to gene-specific criteria and if they matched a pre-specified list of previously reported variants ^13,42^ (Supplementary Table 2). Artifacts were removed as per recommended post-variant call filtering^42^ implemented in the script *whitelist_filter_rscript.R* described by Vlasschaert et al. ^42^ (https://github.com/briansha/Annovar_Whitelist_Filter_WDL) with required resources extracted from Terra Workspaces (https://app.terra.bio/#workspaces/terra-outreach/CHIP-Detection-Mutect2), including NEJM_2017_genes_01262020.txt, CHIP_splice_vars_agb_01262020.txt, CHIP_nonsense_FS_vars_agb_01262020.txt, and CHIP_missense_vars_agb_01262020.txt. The script was cloned and modified to account for minor script errors, including the detection of nucleotide duplication introducing an immediate stop codon without a frameshift (denoted by “*”). A python script was used to perform modifications including for synonymous variants comprising a stop codon which were wrongly tagged by the R script (*e.g.: TET2* X2003X, *PPM1D* X606X), and *JAK2* variant F537_K539delinsL was manually added to the list of CHIP mutations. To minimize artifacts, we excluded variants with a consensus sequencing depth (DP) < 20; insertions or deletions in sites within homopolymer runs (a sequence of 5 identical bases) unless there was a total of 10 or more supporting reads and a VAF>8% for these variants; we excluded previously detected sequencing artifacts TP53:NM_000546:c.1129A>C, TP53:NM_000546:c.215C>G, ASXL1:NM_015338.6:c.708G>C, ASXL1:NM_015338.6:c.2444T>C, and ASXL1:NM_015338.6:c.1934dupG variants with VAF < 0.06); and we excluded variants present in 10 individuals or more (≥0.6%) which were not associated to *TERT* variant rs7705526 or (at p-value ≤ 0.10 by Firth’s logistic regression) (Supplementary Table 3).

### VARIANT DETECTION ASSESSMENT

To assess the performance metrics of the multiplex targeted sequencing panel and bioinformatics pipeline, we used two DNA reference standard samples from Horizon (Horizon Discovery Ltd.), Myeloid DNA (5%) and Tru-Q 0 (100% Wildtype), which were combined at varying dilution ratios to provide expected VAF of 2.5%, 1%, 0.5% and 0.1% for the myeloid sample validated variants at an initial VAF of 5%. The Horizon reference standards are well-characterized, cell line-derived, and control material manufactured under ISO 13485. The Myeloid DNA reference standard contains 22 variants across 19 genes relevant to myeloid cancer (https://horizondiscovery.com/en/reference-standards/products/myeloid-dna-reference-standard) and the Tru-Q 0 reference standard is a well-characterized quality control sample for sequencing assays performance assessment. We obtained from horizondiscovery.com the whole exome sequencing VCF files of these samples based on the Agilent SureSelect Clinical Research Exome v3 (CREV3) with NEXTFLEX™ Pre-& Post-Capture Combo Kit and sequenced on an Illumina NovaSeq 6000 instrument at more than 500x. The variants present in either the Myeloid or the Tru-Q 0 sample within the targeted region of our assay were identified, and CH variants were annotated and identified according to our study’s bioinformatics pipeline. The Myeloid reference sample and the Tru-Q sample had 12 and 5 CH variants respectively, providing 15 unique variants of which 7 corresponded to the Myeloid reference sample’s verified allele frequency variants of 5% VAF (Supplementary Table 4). We performed library preparation and sequencing of the reference samples using our targeted multiplex sequencing panel, and applied variant calling and annotation as described above for 5 replicates for dilutions at VAF 2.5% and 1%, and 4 replicates for dilutions at VAF 0.1% and 0.5%. To assess the positive predictive value (PPV; TP/(TP+FP)) and positive percent agreement (PPA; TP/(TP+FN)) of the assay, we relied on the 5 replicate samples with the dilution providing expected VAF of 2.5% with all 15 variants. When considering all variants at a variant read depth threshold (VD) ≥ 1, the mean PPV and PPA were 8% and 96% respectively, however, restricting to variants with VD ≥ 5 increased the PPV to 64% (Supplementary Table 5.a). For a VAF ≥ 2% and a variant depth threshold VD ≥ 5 as filtering criteria, the mean PPV and PPA were 100% and 75% respectively for all 15 variants (Supplementary Table 5.a). The limit of detection was assessed based on the validated mutations in the HORIZON sample (Supplementary Table 5.b and Supplementary Figure 2). For a variant depth threshold VD ≥ 1 as filtering criteria, the PPA was 52% for VAF of 0.1% and 89% for VAF of 0.5%. When restricting to VD ≥ 5, the PPA was 19% for VAF of 0.1% and 61% for VAF of 0.5%.

### STATISTICAL ANALYSES

Changes in VAF were assessed by using the proportion of variant read counts (VD) over the position-specific read depth (DP) modeled using a generalized mixed regression model fitting a beta-binomial distribution using a logit link function. The beta-binomial model was selected for its ability to directly model VAF while accounting for variability in read depth and overdispersion of the data, and the logit link function ensures that VAF values are bounded in [0,1]. Any variant observed in either the first or end-of-study timepoint sample with a VD of 5 or more was included, along with its matching observation at the other timepoint, including when the matching observation had a VD of 0 or below 5. Multiple variants per patient were accounted for by including the patient as a random factor in the model. The timepoint variable representing the first and end-of-study samples was the exposure of interest and estimates were obtained by contrasting the two timepoint observations using maximum likelihood estimation. Analyses were adjusted for age and the time interval between the two timepoint collection dates. The model’s regression parameters are interpretable as log odds of VAF (VD/DP). The timepoint regression coefficient estimate represents the ratio of the odds of the VAF at end-of-study sample over the odds of the VAF in the first sample. Differences in VAF change between the colchicine and placebo groups were tested by including an interaction term between the timepoint and treatment group variables. The exponent of the interaction term coefficient estimate represents the increase or decrease of the odds of the VAF in the colchicine end-of-study sample compared to the rest. In sensitivity analyses, further adjustment for the delay in time between study drug initiation and the time of the first DNA sample provided consistent results. Sensitivity analyses considering variants with read counts of 1 or above (VD≥1) at one or both timepoints were conducted using a generalized mixed model fitting a negative binomial distribution to model VD and adjusting for DP as an offset. We tested for change in VD by contrasting the end-of-study sample to the first sample observations using Laplace approximation. The expected mean percentage change in mutation counts from the first to the end-of-study sample is obtained by subtracting one from the exponentiated estimate of the timepoint variable. The negative binomial model was selected as it can better account for the zero-inflation that occurs at this lowered VD threshold than the beta-binomial distribution.

For the analysis of clinical outcomes, we used positively adjudicated data according to the intention-to-treat principle. The composite endpoint consisted of the combination of the COLCOT trial primary endpoint (cardiovascular death, resuscitated cardiac arrest, myocardial infarction, stroke, or urgent hospitalization for angina requiring coronary revascularization) to which we added any coronary revascularization events, to increase the number of observed events and potentially increase statistical power. Time to the first occurrence of the composite endpoint was compared between the two treatment groups using the hazard ratio from a Cox proportional hazard model with adjustment for age and sex. Event dates and censoring dates were complete and event-free patients were censored at the date of study completion or at the date of last contact, whichever was the latest. The proportionality of hazards was verified. To account for the multiple occurrences of composite endpoint events within patients, we performed a recurrent event analysis using the Andersen and Gill (AG) model adjusted for age and sex. Individuals were categorized by considering the mutation with the highest VAF on a per-gene basis, allowing the assignation of individuals to VAF groups (VAF > 0; ≥ 0.5%; ≥ 2%; ≥ 10%). All statistical tests were two-sided and conducted at the 0.05 significance level. Statistical analyses were performed using SAS version 9.4.

## DATA AVAILABILITY

The CH-related mutations detected via error-corrected genomic DNA sequencing are provided in Supplementary Table 9. Due to privacy constraints, other data from COLCOT participants are not publicly accessible. However, access to these data may be requested by contacting the COLCOT study committee through Jean-Claude Tardif. All additional data produced or analyzed during this study are included in the present article.

## CODE AVAILABILITY

Source code and guidelines are found in our code repository directory on Github: https://github.com/pgxcentre/COLCOT-CH-scripts. The source code from the R-packages used in this study are freely available online (https://cran.r-project.org/).

## EXTENDED DATA

**Supplementary Table 1a.** Targeted coding regions for sequencing (GRCh37), totaling 35,840 bp.

**Supplementary Table 1b.** Targeted sequencing amplicons (CleanPlex, Paragon) totaling 34,994 bp (GRCh37).

**Supplementary Table 2.** List of variants whitelisted as clonal hematopoietic mutations.

**Supplementary Table 3.** Chip variants, associations with TERT SNV rs7705526 and with age.

**Supplementary Table 4.** CHIP variants from the Myeloid DNA and the Tru-Q 0 reference standards.

**Supplementary Table 5.** Variant detection assessments.

**Supplementary Table 6.** Number of study participants with clonal hematopoiesis variants* at varying thresholds of VAF by age group.

**Supplementary Table 7.** Analysis of change in clonal hematopoiesis variant allele frequency between two sample timepoints.

**Supplementary Table 8.** Cox proportional hazards regression of colchicine versus placebo by groups of clonal hematopoiesis mutation carriers.

**Supplementary Table 9.** Clonal hematopoiesis variants detected

**Supplementary Figure 1.** Flow diagram of study participants

**Supplementary Figure 2:** Observed VAF versus expected VAF for control sample dilutions in which 8 clonal hematopoiesis variants of 5% VAF in the Myeloid sample (Horizon) are considered. Based on 8 validated mutations in the HORIZON sample (includes the 7 mutations listed in Supplementary Table 4, plus ASXL1_W796C which was not whitelisted). There were 5 repeat samples per dilution ratio for VAF 0.025 and 0.01, and 4 repeat samples for VAF 0.001 and 0.005.

## ACKNOWLEDGEMENTS

We thank the Montreal Health Innovations Coordinating Center (MHICC) who managed support activities for the clinical study, all clinical research staff who conducted the study, and all study participants. The study medication and matching placebo were provided by Pharmascience (Montreal), which had no role in the design or conduct of the trial or the preparation or review of the manuscript.

