## Supplementary Figures for "Reduction of clonal hematopoiesis mutation burden in coronary patients treated with low-dose colchicine"

SUPPLEMENTARY MATERIAL

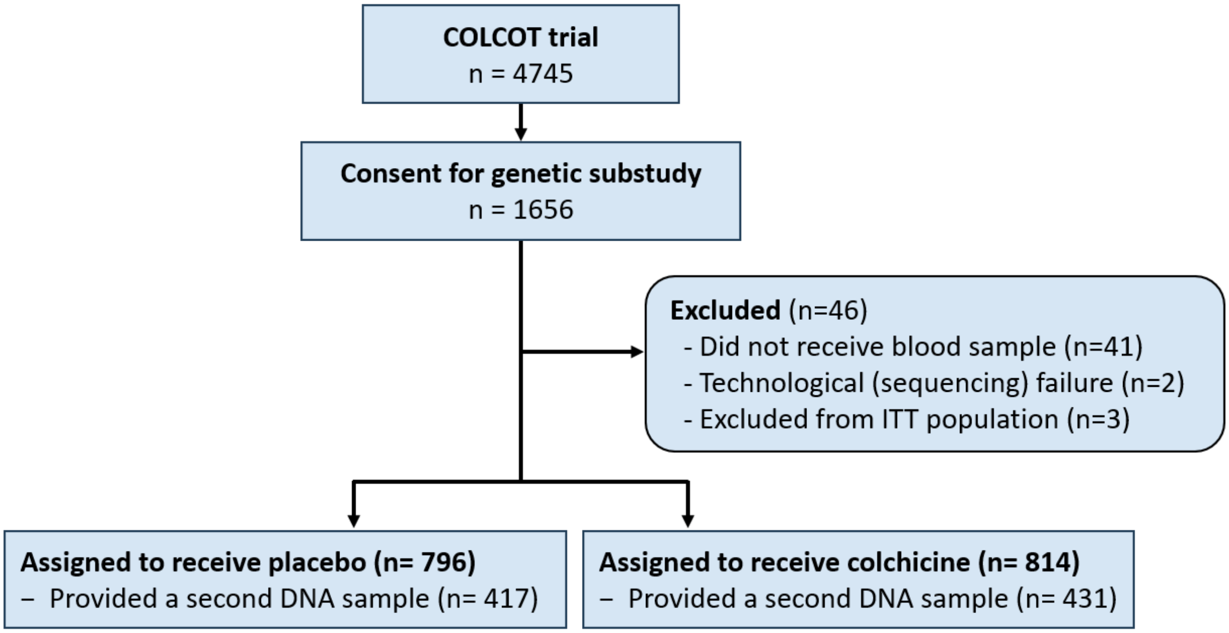


**Supplementary Figure 1.** Flow diagram of study participants

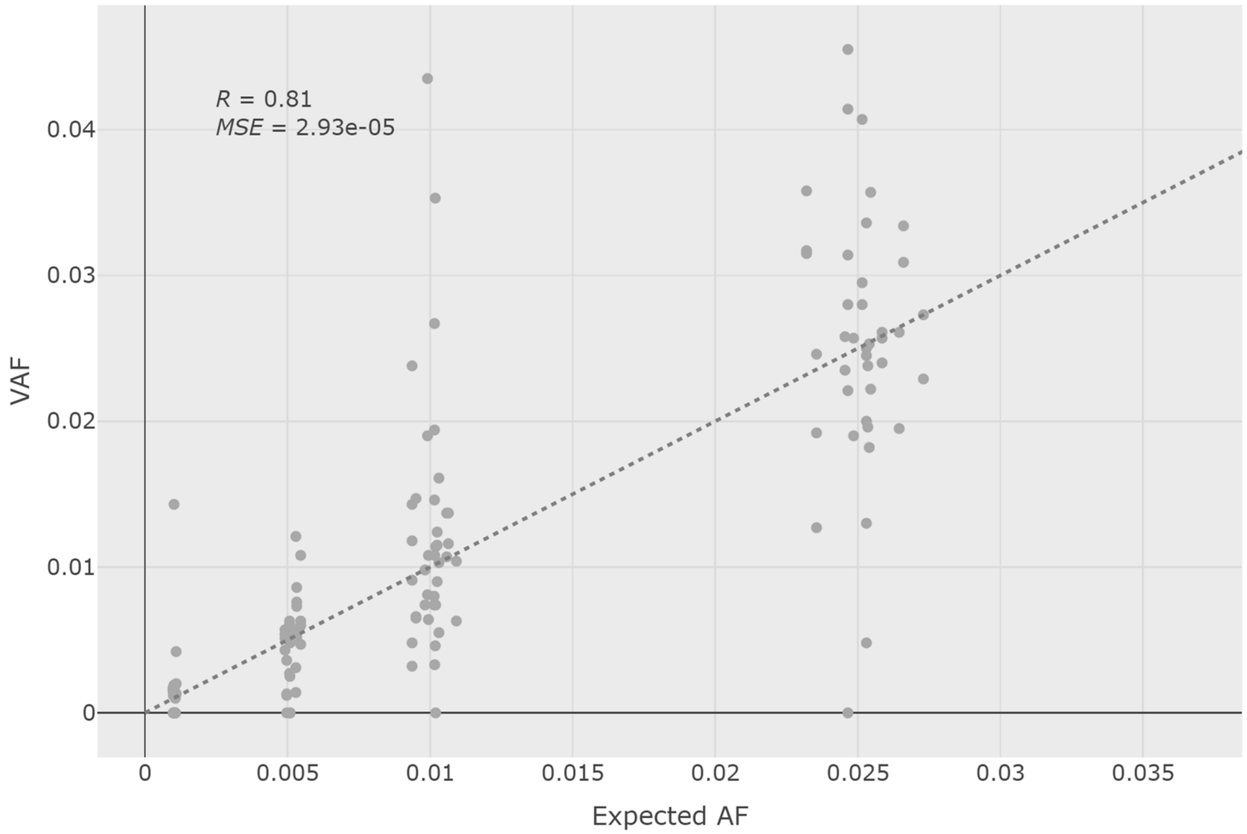


**Supplementary Figure 2:** Observed VAF versus expected VAF for control sample dilutions in which 8 clonal hematopoiesis variants of 5% VAF in the Myeloid sample (Horizon) are considered. Based on 8 validated mutations in the HORIZON sample (includes the 7 mutations listed in Supplementary Table 4, plus ASXL1_W796C which was not whitelisted). There were 5 repeat samples per dilution ratio for VAF 0.025 and 0.01, and 4 repeat samples for VAF 0.001 and 0.005.
